# Counting your chickens before they hatch: improvements in an untreated chronic pain population, beyond regression to the mean and the placebo effect

**DOI:** 10.1101/2023.05.05.23289575

**Authors:** Monica Sean, Alexia Coulombe-Lévêque, William Nadeau, Anne-Catherine Charest, Marylie Martel, Guillaume Léonard, Pascal Tétreault

## Abstract

**Background and aims:** Isolating the effect of an intervention from the natural course and fluctuations of a condition is a challenge in any clinical trial, particularly in the field of pain. Regression to the mean (RTM), wherein extreme scores are more likely to be followed by more average scores, may explain some of those observed fluctuations. However, while this phenomenon is relatively well-known, its effect on outcome measures is rarely quantified, and often only evoked as a potential confound. In this paper, we describe and quantify such symptom fluctuations in a chronic pain population in the absence of treatment, and compare the relative stability of various self-reported outcome measures in untreated chronic low back pain (CLBP) patients and healthy controls (HC).

**Methods:** Twenty-three untreated CLBP patients and 25 HC took part in this observational study, wherein they were asked to complete an array of commonly used questionnaires in pain studies (including the Pain Catastrophizing Scale [PCS], State and Trait Anxiety Inventory [STAI], Central Sensitization Inventory [CSI], Pain Disability Index [PDI], Brief Pain Inventory [BPI] etc.) during each of three visits (V1, V2, V3) at 2-month intervals. Scores at V1 were classified into three subgroups (extremely high, normal and extremely low), based on z-scores. The average delta (Δ=V2-V1) was calculated for each subgroup, for each questionnaire, to describe the evolution of scores over time. This analysis was repeated with the data for V2 and V3.

**Results:** High initial scores were likely to be followed by more average scores; for instance, the “extremely high” subgroup for the PCS (a reputably ‘stable’ questionnaire) had an average decrease of 12/52 from V1 to V2. Participants with “average” initial scores tended to show a small decrease over time, and participants with “extremely low” initial scores tended to remain stable. However, while the pattern of fluctuation in the three subgroups was similar across questionnaires, the magnitude of these fluctuations varied greatly. The STAI and CSI were the most stable questionnaires of all, with even the “extremely high” subgroup showing little or no improvement over time. The least stable questionnaires were the PCS, PDI and BPI.

**Discussion and conclusion:** These pain trajectories in untreated patients cannot be attributable to RTM alone because of their asymmetry, nor to the placebo effect as they occurred in the absence of any intervention. We propose that the observed improvements could be the result of an Effect of Care, wherein participants had meaningful improvements simply from taking part in a study and talking about their pain to benevolent research staff, despite the absence of active or sham treatment. These findings have important clinical ramifications. Beyond simply raising a flag as to the existence (and significance) of Effect of Care, we provide a questionnaire-specific baseline of expected fluctuations based on initial score, against which researchers can compare results from clinical trials when trying to isolate the effect of their intervention.

## Introduction

Chronic pain conditions are highly prevalent and debilitating; as such, many clinical trials are trying to identify treatment approaches^1–7^. However, because pain is a highly subjective and variable phenomenon, it is famously difficult to measure accurately – even more so when it comes to measuring *changes* in pain levels. Indeed, pain levels fluctuate naturally, as they are affected by a wide array of biopsychosocial factors such as sleep, mood, expectations, beliefs, etc. ^8–11^. These factors, like pain, are also often subjective and variable, and difficult to assess – and their impact on pain is similarly challenging to measure with precision^12^.

Clinical trials do their best to quantify the effects of their interventions using the most valid and reliable questionnaires at their disposal ^13, 14^; unfortunately, it remains difficult to isolate the effect of an intervention from the natural course and fluctuations of the condition^15^. This is further complicated by a relatively well-known phenomenon: regression to the mean (RTM)^16^.

RTM is *not* another biopsychosocial factor that *influences* pain levels: it is a *statistical concept* that can – in part – describe, explain, and predict those fluctuations. RTM is based on probability distributions, and states that extreme scores are likely to be followed by less extremes scores that are closer to the individual’s own sampling mean^16^. As such, RTM for pain symptoms depends on two factors: 1) the variability in symptom severity for a given subject (i.e., whether this subject’s symptoms tend to be very constant over time, or whether they fluctuate wildly from one day to the next); and 2) the ‘extremeness’ of the subject’s symptoms at baseline, compared to their own sampling distribution (i.e., how severe their symptoms happened to be on the day of measurement, compared to their average symptom severity)^15^.

RTM is a widely known concept, and it is often mentioned in the discussion section of clinical trial reports, as a possible alternative explanation for observed changes in outcomes over time. However, RTM has rarely been the primary focus of a clinical publication with a chronic pain population. Nevertheless, quantifying RTM – and identifying outcome measures that are intrinsically more susceptible to show RTM – is theoretically possible. For example, on any given questionnaire, RTM would predict that an extreme score (high or low) is likely to be followed by a more average score, regardless of treatment effect; in other words, a high score is expected to be followed by a lower score (and vice versa), while a more average score is expected to remain relatively stable. Complex conditions such as chronic pain are affected by a number of biopsychosocial factors which also have intrinsic variability. Different questionnaires measure these different factors in different ways, such that it is possible that two questionnaires show different RTM for the same subject over the same time period. In other words, a questionnaire measuring a comparatively more fluctuating factor will be more susceptible to RTM.

Quantifying RTM in the questionnaires most often used to assess treatment efficacy in the chronic pain population is of clinical significance for two reasons: 1) It may guide the choice of outcome measures selected at the time of study design; and 2) It may improve result interpretation, helping to differentiate treatment effect from RTM.

The study design required for such RTM assessment requires that patients with chronic pain be assessed using a large array of validated, commonly used questionnaires, at different time points (at least twice, ideally more), with no concomitant intervention taking place outside of usual care. Our team had such an observational study taking place to assess changes in brain structure and functional activity over time in patients with chronic low-back pain (CLBP) and healthy controls (HC). We were therefore able to conduct the RTM analysis presented below as part of this study.

The objectives of this analysis were: 1) to describe and quantify the natural trajectory of questionnaire scores over time, based on initial scores, with a subgoal of determining whether the observed fluctuations were compatible with RTM; and 2) to evaluate and compare the stability of each questionnaire over time, in 23 untreated CLBP and 25 healthy controls.

## Methods

All participants provided written informed consent for their participation into the study. Ethics approval was granted from the institutional review board of the Centre intégré universitaire de santé et de services sociaux de l’Estrie – Centre hospitalier universitaire de Sherbrooke (CIUSSS de l’Estrie – CHUS), Sherbrooke, Canada (file number: 2021-3861). The trial has been registered on Open Science Framework (OSF), under the name “Pilot project on brain and lower back imaging of chronic pain” (registration DOI: https://doi.org/10.17605/OSF.IO/P2Z6Y).

### Participants

Participants were recruited using posters at the CIUSSS de l’Estrie-CHUS, Facebook ads, and by word of mouth. Twenty-seven CLBP patients and 25 HC aged 18 to 75 years old took part in this study. HC were matched with CLBP patients for sex and age.

Specific inclusion criteria for the CLBP were: 1) low back pain (≥ 6 months) with or without pain radiating to the legs or radiating to the neck; 2) average pain intensity of ≥ 3/10 in the 24-hour period before the initial visit; 3) pain primarily localized in the lower back; 4) no history of invasive or aggressive treatment to manage their pain (e.g. corticosteroid infiltration, strong doses of opioids or antidepressants). Specific exclusion criteria for HC were: 1) history of chronic pain; 2) pain at the time of testing; 3) an outstanding painful episode within 3 months of enrollment in the study.

Exclusion criteria for the two populations included: 1) neurological, cardiovascular, or pulmonary disorders; 2) comorbid pain syndrome (i.e., fibromyalgia, osteoarthritis, irritable bowel syndrome, migraine etc.); 3) history of surgical intervention in the back; 4) used of opioids, antidepressants, anticonvulsants, or psychostimulants; 5) a corticosteroid infiltration within the past year; 6) pregnancy (current or planned during the course of the study); 7) inability to read or understand French; 8) contra-indication to Magnetic Resonance Imaging (MRI).

### Study design

The study had an observational longitudinal design. All participants attended three sessions at the Centre de recherche du CHUS, where they completed several questionnaires (reported, analyzed and discussed in the present paper), and provided a saliva sample and underwent brain and lumbar MRI (as part of the larger study). These sessions (V1, V2 and V3) took place at two-month intervals.

### Questionnaires

CLBP patients completed eight questionnaires: 1) the Pain Catastrophizing Scale (PCS), 2) Pain Disability Index (PDI), 3) Brief Pain Inventory (BPI); 4) Pain DETECT; 5) Pain Outcomes Questionnaire (POQ), 6) State-Trait Anxiety Inventory (STAI/S-T), 7) McGill Pain Questionnaire (MPQ), and 8) Central Sensitization Inventory (short form) (CSI).

HC completed only the PCS and the STAI/S-T (two questionnaires applicable to a healthy population).

All questionnaires were completed online using the platform “Research Electronic Data Capture” (REDcap), and are presented in **Table 1**.

**Table 1:** Questionnaires completed by CLBP patients and HC

| Questionnaires | Subscales | Description | Items | Scale | Total score | French validated version |
| --- | --- | --- | --- | --- | --- | --- |
| Pain Catastrophizing Scale (PCS)<sup>13</sup> | - | Degree of catastrophic thoughts (helplessness, magnification, and rumination) | 13 | 5-point Likert-scale (“not at all” to “all the time”) | 0 - 52 | Yes<sup>17</sup> |
| Pain Disability Index (PDI)<sup>18</sup> | - | Ability to perform daily activities (home, social, recreational, occupational, sexual, self-care and life support activities) | 7 | Numerical rating scale (NRS) (0 = no disability to 10 = worst disability) | 0 - 70 | Yes<sup>19</sup> |
| Brief Pain Inventory (BPI) *short form*<sup>14</sup> | Pain severity (BPIs) | Intensity of pain (current, average, least and worst pain in the last 24h) | 4 | NRS (0 = no pain; 10= worst pain imaginable) | 0 - 40 | Yes<sup>20</sup> |
|  | Pain interference (BPIi) | Interference of pain with daily activities (sleeping, walking, mood, etc.) | 7 | NRS (0 = no interference; 10=complete interference) | 0 - 70 | Yes<sup>20</sup> |
| PainDETECT (PD)<sup>21</sup> | PD | Evaluates the presence of neuropathic pain components in patients with back-pain, such as burning sensation, electric shocks etc. | 9 | 7 items rated on a 6-point Likert Scale (0=not at all, 5=very strongly), 1 item based on pain behavior pattern score (-1, 0 or 1) & 1 item based on a radiation score (0 or 2) | 0 - 38 | Yes<sup>21</sup> |
|  | Pain Detect Severity (PDs) | Intensity of pain (current, average in the past 4 weeks, worst in the past 4 weeks) | 3 | NRS (0 = no pain; 10= worst pain imaginable) | 0 - 10 | Yes<sup>21</sup> |
| Pain Outcomes Questionnaire (POQ)<sup>22</sup> | - | Global function (eg. mobility, vitality, affect, daily activities, pain, etc.) | 19 | NRS (0 = less symptoms to 10= more severe symptoms) | 0 - 190 | No (In-house translation) |
| State-Trait Anxiety Inventory (STAI-S/T)<sup>23</sup> | State anxiety (STAI-T) | Current anxiety | 20 | 4-point Likert-scale (“not at all” to “all the time”) | 20 - 80 | Yes<sup>24</sup> |
|  | State anxiety (STAI-S) | General anxiety | 20 | 4-point Likert-scale (“not at all” to “all the time”) | 20 - 80 | Yes<sup>24</sup> |
| McGill Pain Questionnaire (MPQ) *short form*<sup>25</sup> | MPQ | Sensory-affective components of pain | 15 | 4-point Likert scale ("no pain"; to "severe pain") | 0 - 45 | Yes<sup>26</sup> |
|  | MPQ intensity (MPQi) | Pain intensity (previous week) | 1 | 100-point visual analogue scale (left anchor: “no pain”; right anchor: “worst possible pain”) | 0 - 100 | Yes<sup>25</sup> |
| Central Sensitization Inventory (CSI) *short form*<sup>27</sup> | - | Symptoms of central sensitization | 25 | 5-point Likert-scale (0 = “never” ; 4 = “always”) | 0 - 100 | Yes<sup>28</sup> |

To avoid fatigue caused by filling out multiple questionnaires, the PDI (for the CLBP participants) and the PCS (for all participants) were completed at home, one week before each visit. These two questionnaires were chosen because they make no measure of “now” (unlike the other questionnaires), instead measuring more general variables that are unlikely to change over the course of a few days. The remaining questionnaires were completed at the Centre de recherche du CHUS. Questionnaires were always completed in the same order, as listed below.

#### Pain Catastrophizing Scale

The Pain Catastrophizing Scale (PCS) is based on three components of pain catastrophizing: helplessness, magnification, and rumination^13^, and consists of 13 items rated on a five-point likert-scale (“not at all” to “all the time”) with a score ranging from 0 to 52 (where 52 corresponds to high levels of catastrophizing). The French validated version was used^17^.

#### The Pain Disability Index

The Pain Disability Index (PDI) is a questionnaire based on different areas of day living activities such as, home, social, recreational, occupational, sexual, self-care and life support activities, consisting of seven items rated on a numerical scale (0 = no disability to 10= worst disability)^18^. PDI total score ranges from 0-70. The French validated version was used^19^.

#### The Brief Pain Inventory

The Brief Pain Inventory (BPI) short form consists of two subscales, assessing 1) pain severity (BPIs) and 2) pain interference with function (BPIi)^14^. The BPIs assesses current pain intensity, and average, worst and least pain in the last 24 hours. These 4 items are rated on a numerical rating scale (0 = no pain; 10= worst pain imaginable), for a total score between 0 and 40. The BPIi assesses different functional components such as mood, sleep, ability to walk, etc., through 7 items rated on a numerical scale (0 = no interference; 10=complete interference), for a total score between 0 and 70. The French validated version was used^20^.

#### The PainDETECT

The PainDETECT (PD) evaluates the presence of neuropathic pain components in patients with back-pain, such as burning sensation, electric shocks etc.^21^. The PD comprises 7 items related to neuropathic symptoms rated on a 6-point Likert Scale (0=not at all, 5=very strongly), to which a pain behavior pattern score (–1 to 1) and a radiation score (0 or 2) are added, for a total score between 0 and 38.

The PD also includes 3 pain-intensity scores (current pain intensity, average pain in the last 4 weeks, and worst pain in the last 4 weeks), which we averaged into a single score called PDs, ranging from 0 to 10 (0 = no pain; 10 = worst pain imaginable). The French validated version was used^21^.

#### The Pain Outcomes Questionnaire

The Pain Outcomes Questionnaire (POQ) is based on a wide array of components, such as pain, mobility, activities of daily living, vitality, negative affect and fear^22^. The POQ comprises 19 items evaluated on a scale from 0 to 10, for a total score ranging from 0 (least symptoms) to 190 (most severe symptoms). There was no validated French translation available, so we used in-house translation for this questionnaire.

#### The State-Trait Anxiety Inventory

The State-Trait Anxiety Inventory (STAI-S/T) consists in two subscales assessing 1) State (i.e., current) anxiety, and 2) Trait (i.e., general) anxiety respectively^23^. Each subscale comprises 20 items rated on a 4-point Likert scale, for a total score between 20 (very low anxiety) to 80 (very high anxiety). The French validated version was used^24^.

#### The McGill Pain Questionnaire

The McGill Pain Questionnaire (MPQ) short form evaluates the sensory-affective components of pain using 15 items evaluated on a 4-point Likert Scale (0=no pain; 3=severe pain), for a total score ranging from 0 to 45^25^.

The MPQ also includes a separate question assessing the average pain intensity over the previous week, using a 100-point visual analogue scale (left anchor: “no pain”; right anchor: “worst possible pain”). This score was called MPQi and was analyzed separately from the rest of the MPQ. The French validated version was used^26^.

#### The Central Sensitization Inventory

The Central Sensitization Inventory (CSI) short form assesses symptoms suggesting the presence of central sensitization or central sensitivity syndromes, using is 25 items rated on a five-point Likert-scale (0=“never”; 4= “always”)^27^, with total scores ranging from 0 to 100. The French validated version was used^28^.

### Data Processing and Statistical Analysis

#### Group attribution

To first describe the behavior of “extreme” vs “normal” scores, it was necessary to establish a criterion to differentiate “extreme” and “normal” scores. This was done by transforming initial raw scores into studentized scores (i.e., z-scores) for each questionnaire. Multiple z-score thresholds were tested, and a threshold of |z| > 0.5 was found to yield the most similar number of participants across the 3 subgroups (“extremely high”, “normal”, “extremely low”). As such, scores with |z| > 0.5 (i.e., scores that were more than half a standard deviation above or below the group average) were considered “extreme”, whereas scores with |z| < 0.5 (i.e., scores within half a standard deviation of the group average) were considered “normal”. An exploratory analysis with various thresholds revealed that, regardless of the threshold used, a similar pattern emerged from our results. All results obtained using the different thresholds tested (|z| > 0.66; |z| > 0,8 and |z| > 1) are included in **supplementary materials**.

Scores at V1 were thus classified as a) extremely high, b) normal, or c) extremely low. This was done independently for each questionnaire, such that a given participant could be in the “extremely high” subgroup for one questionnaire, but in the “normal” subgroup for another questionnaire. Next, the delta between V1 and V2 was calculated by subtracting the score at V1 from the score at V2 (Δ=V2-V1), such that a positive delta corresponds to a score increase (i.e., worsening of the condition), and a negative delta corresponds to a score decrease (i.e., improvement of the condition). The same analysis was conducted between V2 and V3: scores at V2 were again classified as a) extremely high, b) normal, or c) extremely low, and the delta between V2 and V3 was calculated by subtracting the score at V2 from the score at V3 (Δ=V3-V2). Thus, for each questionnaire, two calculations were performed (V2-V1 and V2-V3).

#### Standardization across questionnaires

To facilitate the comparison between questionnaires, which use various scales, all raw scores were reported on a scale from 0 to 100. Fluctuations larger than 10 percentage points between visits were considered clinically meaningful, and fluctuations of 5 percentage points or less were considered random noise. Fluctuations between 5 and 10 percentage points (5 |Δ| ≤10), while of debatable clinical relevance, were still considered likely enough to denote an effect to warrant being reported. The use of such standardized thresholds, as opposed to the Minimal Detectable Change (MDC) specific to each questionnaire, was favored because it allowed for direct comparisons between questionnaires; the advantages and drawbacks of this methodological choice are highlighted in the Discussion.

#### Average delta scores

Once participants were divided into the three subgroups (based on their initial scores), average delta scores were calculated for each subgroup within each questionnaire. This yielded a measure of the average evolution over time for each subgroup.

#### Fluctuation scores

In addition to *average* delta scores, “fluctuation scores” were calculated for each subgroup within each questionnaire by averaging the *absolute value* of delta scores for the given subgroup. This yielded a measure of the magnitudes of fluctuations at play, regardless of their direction. This measure was particularly informative in cases where both large decreases and large increases had taken place. For example, a questionnaire could have an average score for all participants of 27/100 at V1 and of 28/100 at V2, yielding an *average* delta score of only 1/100 – seemingly very stable over time. However, such ‘stability’ could actually be the result of large increases in some participants and large decreases in others, cancelling each other out. If that were the case, while the *average* delta would be close to 0, the *fluctuation* score would be large, thus more accurately representing the variability of scores over time for that particular questionnaire.

#### Inferential statistics

In line with the exploratory nature of this paper and given the small sample size, emphasis was placed on descriptive statistics.

## Results

### Participants

Fifty-two participants (25 HC and 27 CLBP) were recruited in the study. Three CLBP participants dropped out after the first visit (unexpected pregnancy [n=1], discomfort during MRI [n=1], scheduling conflicts [n=1]), and one dropped out after the second visit (move to a different city [n=1]), such that 23 CLBP completed the entire study and were included in the analysis. There were no dropouts among the HC.

The 25 HC (11 women, 12 men) were aged 44 ± 15 years old, and the 23 CLBP participants (15 women, 11 men) were aged 40 ± 14 years old. Other sociodemographic characteristics of the sample are presented in **Table 2**. All participants complied with the instructions relating to medication/treatment throughout the study, namely, that they were to avoid any treatment other than over-the-counter medication and their usual, non-invasive rehabilitation treatments. This allowed us to evaluate the natural course of the condition during the period of the study.

**Table 2:** Sociodemographic characteristics of the sample.

|  | CLBP (n=23) | HC (n=25) |
| --- | --- | --- |
| **Biological sex** |  |  |
| Women | 11 | 15 |
| Men | 12 | 10 |
| **Age** (average ± sd) | 44 ± 15 | 40 ± 14 |
| **Ethnicity** |  |  |
| Caucasian | 16 | 24 |
| Asiatic | 1 | 1 |
| Hispanic | 4 | - |
| African | 1 | - |
| Arabic | 1 | - |
| **Education level** |  |  |
| Primary school | - | - |
| High school | - | - |
| Apprenticeship | 5 | 2 |
| College | 4 | 6 |
| University | 14 | 17 |
| **Annual income** |  |  |
| less than 20K | 2 | 5 |
| 20K - 35K | 5 | 2 |
| 35K - 50K | 5 | 1 |
| 50K - 65K | 5 | 6 |
| 65K - 80K | 2 | 5 |
| 80K - 100K | 3 | 4 |
| 100K and more | 1 | 2 |
| **Pain duration** |  |  |
| 4 month - 5 months | 0 | NA |
| 6 months - 12 month | 5 | NA |
| 1 - 4 years | 7 | NA |
| 5 years and more | 11 | NA |

Participants completed each questionnaire 3 times throughout the study, with 2 months between each visit. The average scores for each questionnaire at each visit are presented in **Table 3a and 3b** for the two populations. The evolution over time of ‘extremely high’ scorers, ‘extremely low’ scorers, and ‘normal’ scorers in the CLBP sample (see *Statistical Analysis* for a detailed description of the analysis method) is shown in Table 4, represented in 3 different ways (Table 4a, 4b, and 4c). Table 4a shows the *average* evolution for each subgroup. For example, on the PCS, the CLBP participants in the ‘extremely high’ subgroup at V1 (i.e., participants with a z-score >0.5) had, on average, a reduction of 22 percentage points in their scores at V2 – corresponding to a reduction of 12 points on the PCS scale (range: 0-52). Table 4b shows the same data but with *individual* delta scores as opposed to *average* delta scores for each subgroup. For example, for the PCS, the top left cell shows all individual delta scores (from V1 to V2) for the participants classified in the ‘extremely high’ subgroups at V1: two participants had a reduction in score ∼30 percentage point, and four participants had a reduction in score ∼20 percentage points. These individual scores, averaged together, yield the average score (–22) reported in table 4a. Table 4c shows the ‘fluctuation scores’, corresponding to the average of the *absolute value* of the deltas for each subgroup (in the case of the PCS scores from V1 to V2, for the ‘extremely high’ subgroup, this fluctuation score is equal to the mean delta score because all deltas were negative; however, for the ‘normal’ subgroup, the fluctuation is score is substantially larger than the average delta score, because of the presence of both positive and negative delta scores).

**Table 3:**
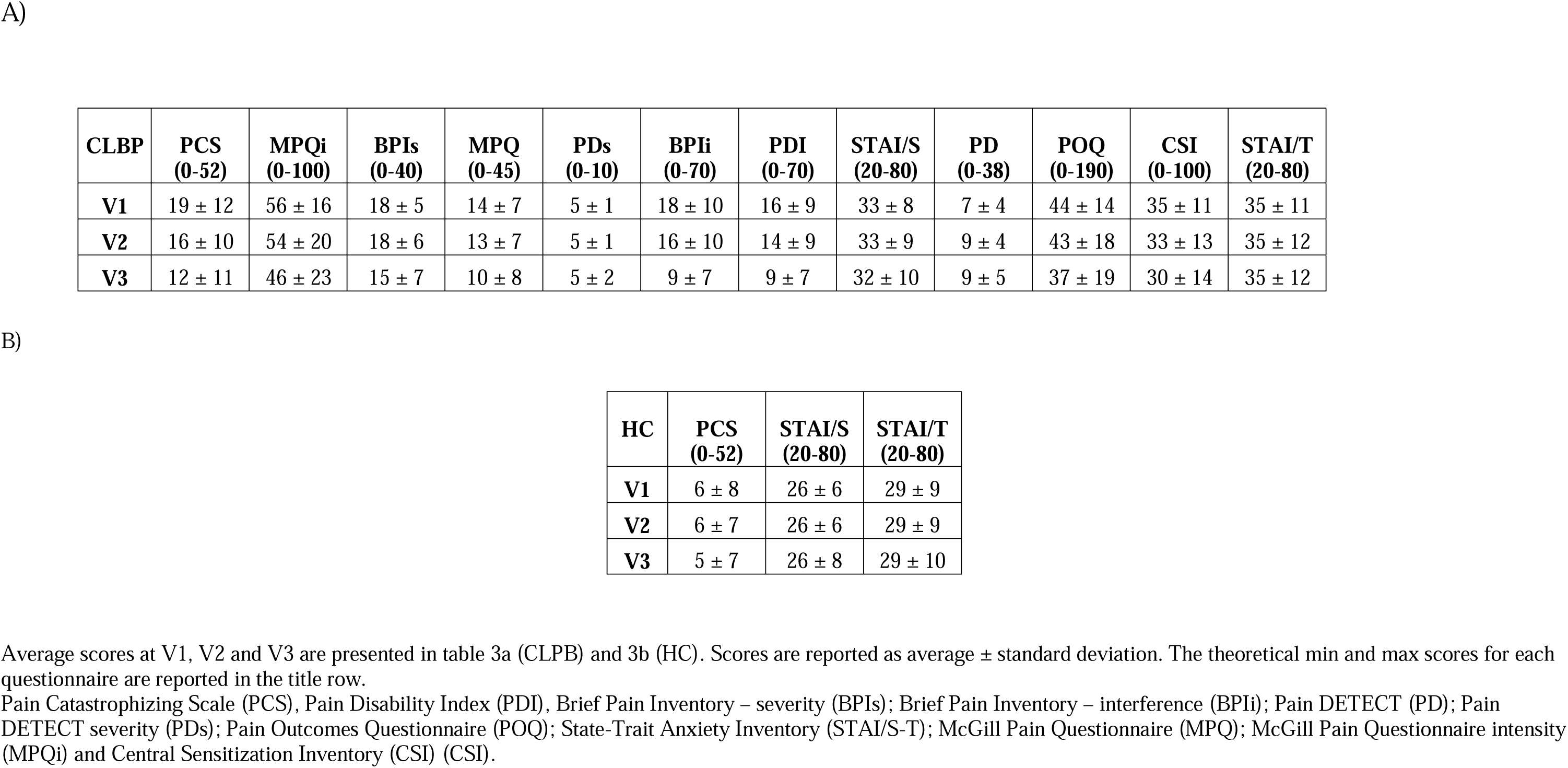
Average scores for each questionnaire during the 3 visits (V1, V2, and V3), for the CLBP sample and the HC sample.

**Table 4:**
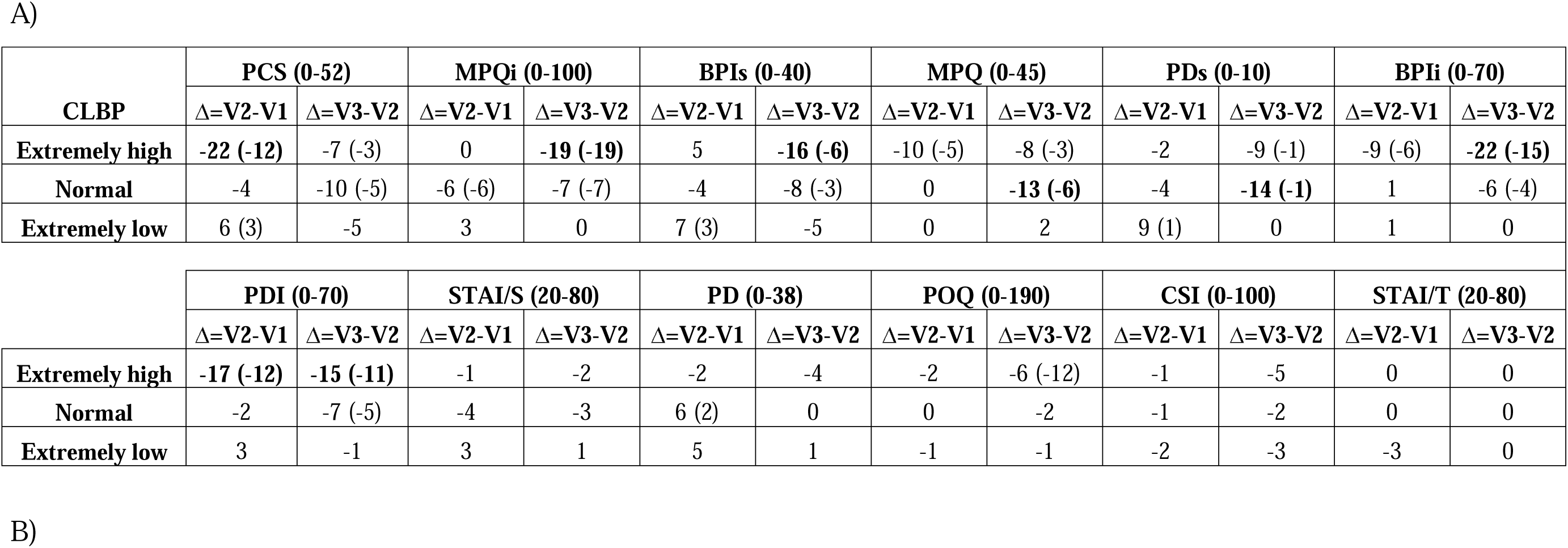
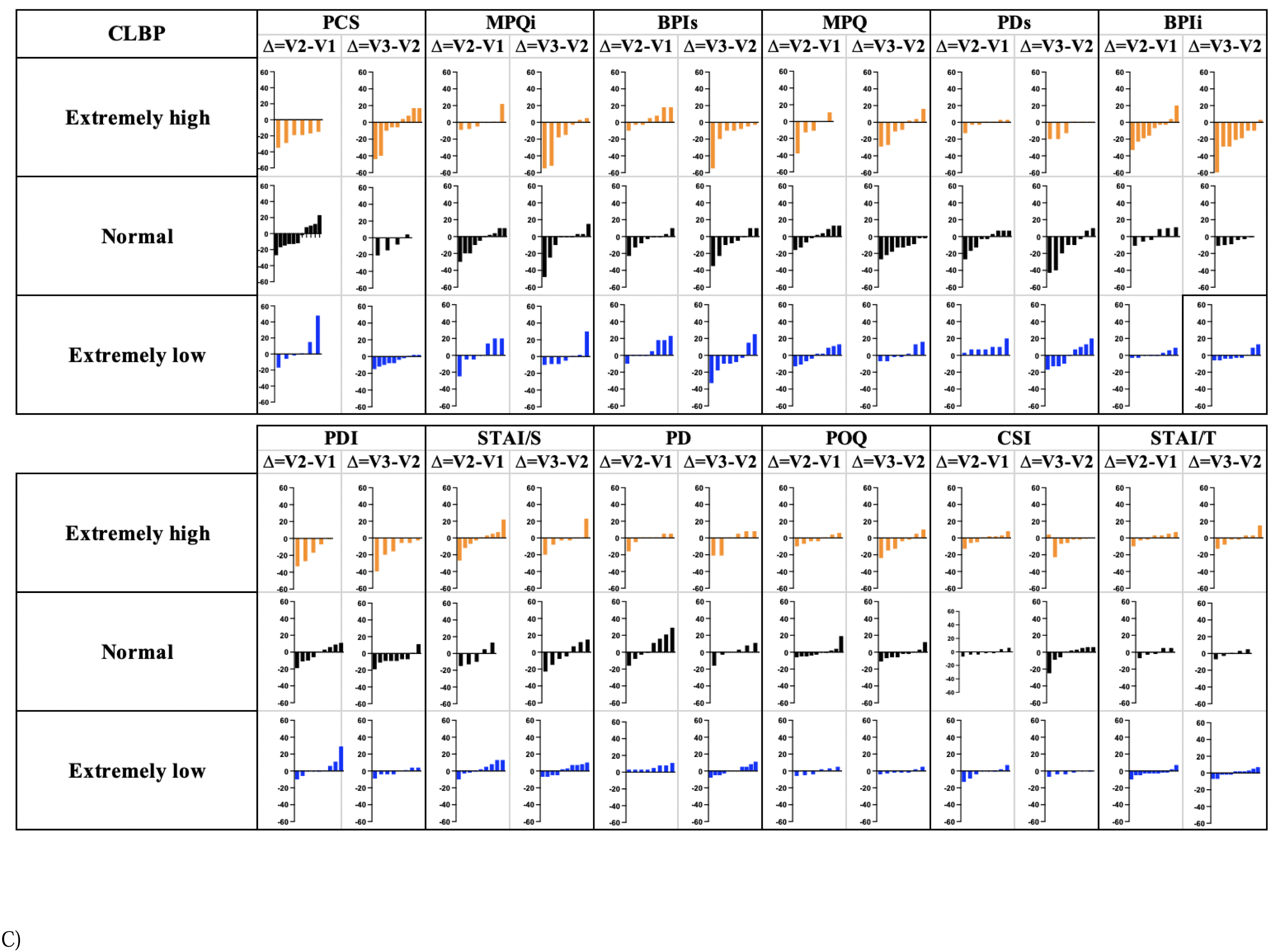
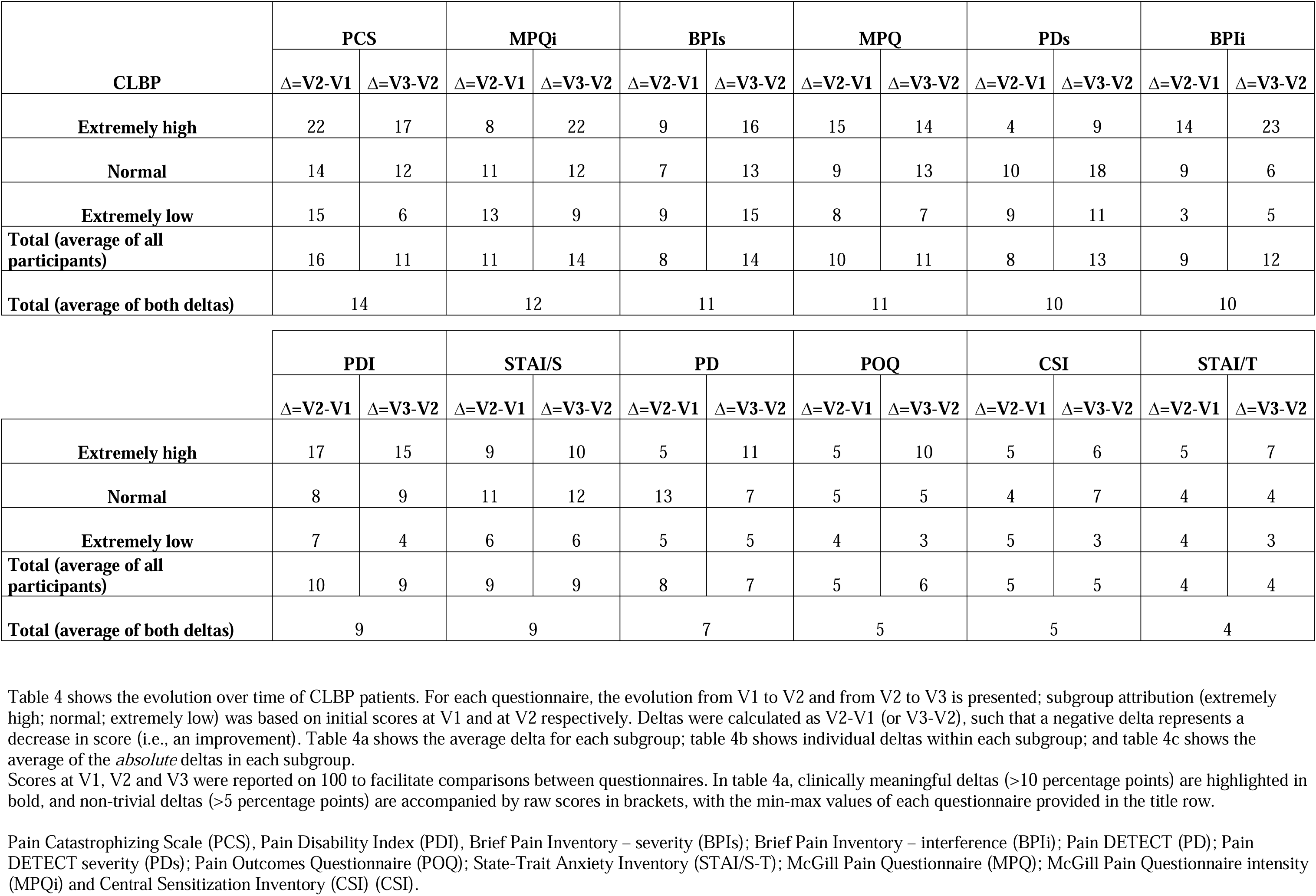
Evolution over time based on initial score – CLBP sample.

As stated in the methods, deltas smaller than or equal to 5 percentage points were considered random noise and most likely not denoting a real change, while deltas larger 10 percentage point were considered clinically meaningful. Deltas between 5 and 10, while arguably not clinically meaningful, were still considered likely to be more than simple noise, and therefore to be worth reporting.

### Average evolution over time as a function of initial score

#### Average evolution of ‘extremely high’ scores

As shown in the top rows of Table 4a and 4b, participants with an initial ‘extremely high’ score at V1 tended to show a reduction in score at V2, and those with an ‘extremely high’ score at V2 similarly tended to show a reduction in score at V3. Indeed, the *average* deltas from V1 to V2 and from V2 to V3 were mostly negative in that subgroup (table 4a, top row) and most *individual* deltas were negative (table 4b, top row). This trend for high scores to be followed by lower scores is expected, as RTM predicts that extreme scores will be followed by more normal scores, which in the case of extremely *high* scores means that the subsequent score should be *lower*.

#### Average evolution of ‘normal’ scores

The evolution of ‘normal’ scorers is shown in the second rows of Table 4a, 4b and 4c. As shown in table 4a, *average* scores tended to remain stable or to slightly decrease over time. Indeed, out of the 24 average delta scores obtained in this subgroup (table 4a, row 2), 15 were negligible (<5 percentage points). Out of the remaining 9 data points, which are all negative, only two show an average decrease large enough to be considered clinically meaningful (one instance in the MPQ, and the other in the PDs).

The visual representation of individual delta scores (Table 4b, middle row) reveals that participants in this subgroup tended to show an uneven split between increases and decreases in scores from one visit to the next, with a larger number of individual delta scores being negative.

RTM predicts that participants with a ‘normal’ score will show little or no change from one session to the next, with an even distribution of increases and decreases cancelling each other out. This should translate in average delta scores being roughly equal to 0 (Table 4a) and, visually, in a roughly even and symmetrical split between individual increases and decreases (Table 4b). As this is not the pattern of results that we observed, our results suggest that RTM alone cannot account for the overall decrease in scores seen in some outcome measures (see discussion).

#### Average evolution of ‘extremely small’ scores

The evolution of ‘extremely low’ scorers (i.e., participants with an initial Z score <-0.5 at V1 for the V1-V2 analysis, and those with a Z score <-0.5 at V2 for the V2-V3 analysis) is shown in the third rows of Table 4a, 4b and 4c. As it can be seen in table 4a, on average, this subgroup appears to remain stable from one visit to the next on most questionnaires, with only sparse and small average increases observed on a few questionnaires. However, on roughly half of the questionnaires, these seemingly ‘stable’ average deltas are a product of significant individual increases and decreases that roughly cancel each other out, as shown visually in table 4b and quantified in table 4c. In the other questionnaires, the average stability does appear to stem from a widespread absence of individual fluctuations, as evidenced by the data presented in table 4b and 4c.

RTM predicts that extremely low scores will *increase* towards more ‘normal’ scores on the subsequent measurement. Overall, there appears to have been a slight RTM effect on a few questionnaires, although most average deltas are close to 0, suggesting that no substantial RTM was at play – or that some other effect was at play that counteracted RTM (see discussion).

### Analysis by questionnaire

#### Average evolution

For the ‘extremely high’ subgroup, an overall average decrease in score was observed from one visit to the next. However, this effect was stronger in certain questionnaires: notably, the PCS, PDI, BPIi, BPIs, and MPQi all showed an average decrease larger than 15 percentage points for at least one time period (Table 4b). On the other hand, other questionnaires remained comparatively more stable: the PD, CSI, and both subscales of the STAI showed no change on average from one visit to the next, for both time periods.

The ‘normal’ subgroup was more stable overall, with clinically meaningful average fluctuations (as we defined: Δ<-10) observed only for the MPQ and PDs, over a single time period. As with the ‘extremely high’ subgroup, the ‘normal’ subgroup was most stable on average on the CSI and both subscales of the STAI, as well as on the POQ (table 4b).

The ‘extremely low’ subgroup had the most stable scores of all, showing no clinically meaningful average change on any questionnaires, and only showing small (between 5 and 10 percentage points) average changes on the PCS, PDs and BPIs, for a single time period (table 4b).

Overall, the PCS shows the largest average change for all three subgroups, followed by the PDI, PDs, both subscales of the MPQ, and both subscales of the BPI. The questionnaires with the smallest average delta were the CSI and both subscales of the STAI (table 4b).

#### Average absolute fluctuation

As mentioned previously, it is possible for a questionnaire to have an average delta of roughly 0 from one visit to the next, seemingly suggesting that all participants remained stable over time, while in fact large individual increases and decreases in scores have been taking place, cancelling each other out. Fluctuations scores (table 4c), computed by averaging the absolute value of individual deltas within each subgroup and questionnaire, allows us to identify such cases. For example, on the PCS, the ‘normal’ subgroup had an average delta of –4 between V1 and V2. At face value, this suggests that scores remained roughly unchanged between V1 and V2 for participants in this subgroup. However, visual inspection of table 4b shows that significant increases and decreases seem to have taken place, and taking the average of the *absolute value* of these deltas allows us to quantify the average magnitude of the fluctuations – in this case, 14 percentage points (table 4c). This suggests that even for a participant whose initial score on the PCS was ‘normal’ (i.e., not extreme), a fairly large change in score can be expected at the following visit. In contrast, for the ‘extremely low’ subgroup in the STAI-T – which has an equally small average delta of 3 percentage points (table 4a) – the average of the absolute value of these deltas is 4, suggesting that this questionnaire is truly stable.

For all three subgroups, the PCS shows the largest fluctuation in scores, followed closely by the MPQi, BPIs, MPQ, PDs, and BPIi. The STAI-T was the only questionnaire where participants had, on average, fluctuations smaller than 5 percentage points across visits.

#### Healthy controls

HC completed the same visits and procedures as the CLBP patients but completed only the questionnaires applicable to healthy subjects: the PCS and both subscales of the STAI.

HC had much more stable scores overall compared to CLBP patients (Table 5). Indeed, the average evolution over time is smaller than 5 percentage points for all 3 subgroups, on all questionnaires and at both time points (with one exception at 6 percentage points) (Table 5a). Visually, apart from a notable outlier on the STAI-S (a grad student who reported having a particularly stressful day), most individual fluctuations from one visit to the next were also negligible (Table 5b). This is further supported by the average *absolute* fluctuation scores, which for the most part do not meet the threshold to be considered clinically meaningful (Table 5c). The only exception appears to be the ‘extremely high’ subgroup for the PCS, with average *absolute* fluctuations of 11 and 14 percentage points from V1 to V2 and V2 to V3 (respectively). However, it is important to point out that this subgroup was comprised of only 3 participants.

**Table 5:** Evolution over time based on initial score – HC sample.

A)
| HC | PCS (0-52) |  | STAI/S (20-80) |  | STAI/T (20-80) |  |
| --- | --- | --- | --- | --- | --- | --- |
| | $\Delta=V2-V1$ | $\Delta=V3-V2$ | $\Delta=V2-V1$ | $\Delta=V3-V2$ | $\Delta=V2-V1$ | $\Delta=V3-V2$ |
| Extremely high | -4 | 1 | -1 | -6 (-4) | -2 | 3 |
| Normal | -1 | -4 | 0 | 2 | 1 | -5 |
| Extremely low | 1 | 0 | 2 | 2 | -1 | 3 |

| HC | PCS |  | STAI/S |  | STAI/T |  |
| --- | --- | --- | --- | --- | --- | --- |
| | $\Delta=V2-V1$ | $\Delta=V3-V2$ | $\Delta=V2-V1$ | $\Delta=V3-V2$ | $\Delta=V2-V1$ | $\Delta=V3-V2$ |
| **Extremely high** | 11 | 14 | 5 | 8 | 8 | 5 |
| **Normal** | 5 | 5 | 8 | 11 | 5 | 6 |
| **Extremely low** | 3 | 1 | 3 | 2 | 1 | 4 |
| **Total (average of all participants)** | 5 | 5 | 5 | 7 | 4 | 5 |
| **Total (average of both deltas)** | 5 |  | 6 |  | 5 |  |

Scores at V1, V2 and V3 were reported on 100 to facilitate comparisons between questionnaires. In table 5a, clinically meaningful deltas (>10 percentage points) are highlighted in bold, and non-trivial deltas (>5 percentage points) are accompanied by raw scores in brackets, with the min-max values of each questionnaire provided in the title row.
Pain Catastrophizing Scale (PCS), and State-Trait Anxiety Inventory (STAI/S-T)

Interestingly, all three questionnaires completed by the HC (the PCS, STAI-S, and STAI-T) had an average absolute fluctuation of roughly 5 percentage point across all subgroups and both time points (Table 5c). In contrast, those same questionnaires in the CLBP population had an average absolute fluctuation of 14, 9 and 4, respectively (Table 4c).

## Discussion

The objectives of this analysis were: 1) to describe and quantify the natural trajectory of questionnaire scores over time, based on initial scores, with a subgoal of determining whether the observed fluctuations were compatible with RTM; and 2) to evaluate and compare the stability of each questionnaire over time, in 23 untreated CLBP and 25 healthy controls.

Our results show that the CLBP population had relatively large variations in outcome measures over time, and that this effect varied across subgroups and across questionnaires. It bears repeating that these fluctuations were *observed in the absence of any experimental intervention*. Participants with high initial scores were overwhelmingly likely to show a decrease in score at the subsequent measurement, while participants with normal or extremely low scores were relatively more stable. In terms of questionnaires, the PCS showed the most variation in scores over time; both subscales of the MPQ and both subscales of the BPI as well as the PDs also showed meaningful variations. The most stable questionnaire overall was the STAI-T, followed by the CSI and POQ. Healthy controls, in contrast, showed very little variability. Average deltas and average *absolute* deltas were similar – and very small – across all subgroups and questionnaires.

RTM alone cannot be responsible for all the variability observed in our CLPB sample. Indeed, RTM predicts that extreme scores are likely to be followed by less extremes (more ‘normal’) scores; as such, if RTM was the sole driving factor, extremely high scores would be followed by lower scores, and extremely low scores would similarly be followed by higher scores. However, while we did observe that extremely high scores were generally followed by lower scores, extremely low scores were *not* followed by higher scores, instead remaining relatively stable. Moreover, RTM predicts that ‘normal’ scores are equally likely to show a slight increase or a slight decrease at the subsequent measurements, and that these random variations should roughly cancel out. As such, if RTM was the sole driving factor, the average delta in the ‘normal’ subgroups should be roughly 0. However, what we observed was a tendency for scores in the ‘normal’ subgroup to *decrease* over time.

Together, these results suggest the presence of an effect responsible for a generalized decrease in scores (i.e., clinical improvement) over time. We hypothesize that this effect is a result of the attention and care received by the patients as part of their participation in the study, and as such propose calling this effect “Effect of Care”. Indeed, even if a participant is fully aware that they are not receiving any treatment (which therefore rules out a placebo effect, in its textbook definition^29^), simply having the chance to talk about their pain with understanding, thoughtful and competent-looking research staff could contribute to improve their symptoms. Additionally, the ‘seriousness’ afforded by the inclusion of brain and lumbar MRI – a notably well-regarded and imposing modality – likely further increased the potency of Effect of Care in our study.

### Similarities and differences with Test-Retest

At first glance, this study presents superficial similarities with the well-known test-retest, such that readers might question the novelty and relevance of our findings. However, our analysis presents a number of significant differences with test-retest, and as such can offer novel and clinically relevant findings.

First, test-retest is often included as part of a validation study for a single questionnaire. Therefore, despite efforts to standardize these studies and improve generalizability, the varying study design and populations can make it difficult to compare different questionnaires. In the present study, a single patient population completed a large array of questionnaires within a fixed time frame. As such, we can easily isolate and compare the variability attributable specifically to the questionnaires.

Moreover, while test-retest studies generate a single overall score for a questionnaire, we conducted an analysis by subgroup. This allowed us to isolate and quantify differing degrees of variability *within* a questionnaire, and to highlight directional trends depending on the initial score, providing more nuanced and precise results than a single overall score.

### Biases and limitations

The most important limitation in this study is obviously the small sample size, especially as we further divided our sample into three subgroups. These subgroups were also of varying sizes, a result of our method of obtaining these subgroups, based on z-scores. However, having three assessment time points allowed us to conduct two separate analyses (V1 to V2, and V2 to V3) which showed similar results. Furthermore, the objective of this study was not to precisely quantify specific effects, but rather to explore a data set and identify general trends and effects. Finally, the fact that a similar pattern was found regardless of the classification threshold used (see supplementary materials) lends further credibility to our findings.

Another potentially objectionable point was the decision to use an arbitrary threshold for fluctuations that are considered ‘noise’ (≤5/100) vs ‘clinically meaningful’ (>10/100), as opposed to using the established Minimal Detectable Change (MDC) or Minimal Clinically Important Difference (MCID) of each instrument. Standardized thresholds were chosen to facilitate comparisons between questionnaires, which would otherwise have been counterintuitive at best. This decision was again consistent with our objectives, which were to identify overall trends and not to quantify phenomena with a high degree of precision (which would have been impossible given our small sample size).

### Relevance for clinical trials

It is difficult to determine with certainty whether the variation in scores observed in this study are a manifestation of RTM or the result of Effect of Care or some other effect. However, being able to quantify this variability for specific questionnaires and specific subgroups has important clinical implications. Indeed, like MDC (which, unlike our, it will allow future researchers conducting clinical trials to compare their observed variations against our results, in order to isolate and better estimate the ‘true’ effect of their intervention). For example, a researcher might be thrilled to see a reduction of 10 points on the PCS following an experimental treatment. However, as shown in our study, such a decrease can easily be observed in the absence of any treatment.

## Conclusion

Ours results showed that CLBP patients with more severe symptoms at baseline will tend to show improvement at the subsequent measurement, even in the absence of an intervention – which could lead researchers to overestimate the effect of their intervention. However, patients with less severe symptoms at baseline do not show the corresponding exacerbation predicted by RTM, and patients with more average symptoms at baseline also tend to show an improvement at the subsequent measurement. These results suggest the presence of an Effect of Care, wherein patients generally show an improvement in symptoms simply by being part of a study.

Our results also provide a preliminary quantification of the variability in scores observed over time, in the absence of an intervention, in a CLBP population. This variability depends on initial score and is different across questionnaires. Our results can therefore be used to guide interpretation of results obtained in clinical trials.

## Data Availability

All data produced in the present work are contained in the manuscript

https://osf.io/p2z6y

