## Supplemental materials for "Counting your chickens before they hatch: improvements in an untreated chronic pain population, beyond regression to the mean and the placebo effect"

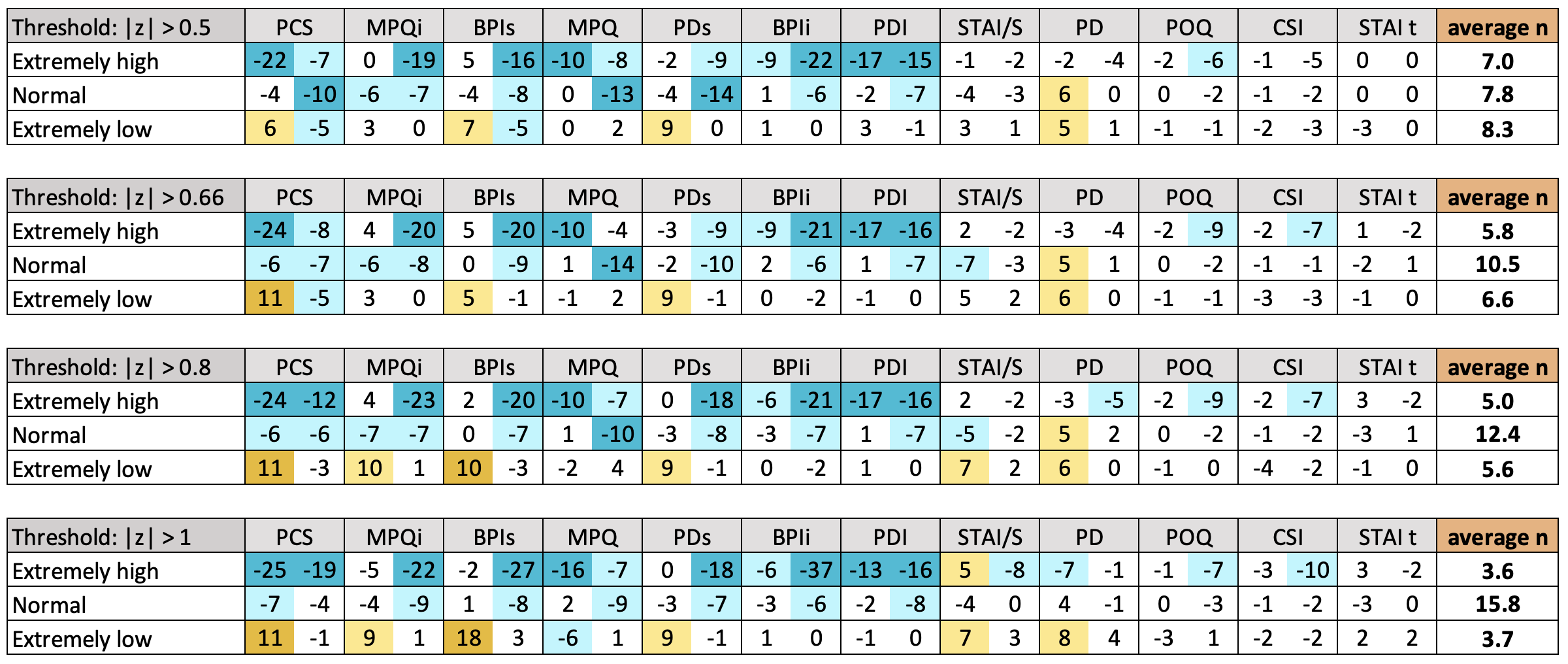

Supplementary Materials

‘Extremeness’ of scores was determined based on z-scores. Different thresholds were tested; the threshold of |z|>.5 was retained as it yielded 3 subgroups of similar sizes, with little effect on overall trends.

This table presents the main results obtained with all thresholds tested (0.5, 0.66, 0.8, 1).

For each questionnaire and each subgroup, the average delta from V1 to V2 is presented in the left column, and the average delta from V2 to V3 in the right column, wherein a negative score represents a reduction in symptom intensity.

To facilitate comparisons across questionnaires, all scores were reported on 100.

Dark blue cells represent average *reductions* larger than 10 percentage points; light blue cells represent average *reductions* between 5 and 10 percentage points; light yellow cells represent average *increases* between 5 and 10 percentage points; dark yellow cells represent average *increases* larger than 10 percentage points.

While exact results vary somewhat depending on threshold (which is not unexpected given that group sizes for the extreme subgroup decreases sharply, increasing the weight of outliers), the same trends appear regardless of thresholds, as highlighted by the color patterns.
